## Supplementary material for "Modeling the impact of xenointoxication in dogs to halt *Trypanosoma cruzi* transmission": S7 Code

Chagas flurolaner model


### Chagas flurolaner model

###### B. Raynor

#### 3/14/2022

Load packages

```
rm(list = ls())

library(deSolve)
library(ggplot2)
library(dplyr)
```

#### Models

##### I. Model One: Bugs and Dogs transmission dynamics pre-fluralaner tx:

```
  - jr added in dm to see how model 2 would look prior to treatment and determine when X and Y reach equilibrium  
  - X doesn't reach equilibrium until after 10000 days  
  - Model one and has been combined with model 2, just here for reference.
```

**Parameters**  
Y is the proportion of triatomines infected  
X is the proportion of dogs infected  
a is expected number of bites on dogs per triatomine (1/14)  
m is equilibrium triatomine density per dog  
n is length of incubation period in insects (45)  
b is the transmission efficiency from infectious triatomines to susceptible dog, through biting (0.00068)  
c is the probability of infection of an uninfected triatomine by biting an infectious dog (0.49, or 0.28- halfed to account for cyclic parasitemia)  
r is daily force of infected dog mortality 1/(365\*3)=0.0009  
g is the daily probability of bug mortality

**baseline, pre-treatment model**

```
#model
RM <- function(time, state, parameters) 
{
  with(
    as.list(c(state, parameters)), 
    {
      dX <- m*a*b*Y*(1-X)-r*X   
      dY <- a*c*X*(exp(-g*n)-Y)-(g*Y)
      dm <- (R*(1-m/K)*m )
      return(list(c(dX, dY, dm)))
    }
  )
}
```

##### II. Model Two: Bugs and Dogs transmission dynamics after fluralaner tx

Extract equilibrium values

```
getEq<-function(a, b, m, n, g, c, r)
{
  R0 <- (m*(a^2)*b*c*exp(-g*n)) / (r*g)
  S<-a/g
  Xeq<-(R0-1)/(R0+(c*S))
  Yeq<-((a*c*Xeq)/(g+a*c*Xeq))*exp((-g*n))
  togo<-c(R0,Xeq,Yeq)
  return(c(R0, Xeq, Yeq))
}

# RMequil <- getEq(a=1/14, b=.00068, m=40, n=45, g=.005, c=.28, r=1/(3*365))
# 
# Xeq <- RMequil[2]
# Yeq <- RMequil[3]
```

estimate parameter z, percent of bugs that die after feeding on treated dogs, from data in Laino et al. 2019 https://www.ncbi.nlm.nih.gov/pubmed/30981313  
x = days post fluralaner treatment  
y = percentage of bugs that died after feeding on treated dogs (5th stage pyrethroid resistant instars) NOT USED  
z = percentage of bugs that died after feeding on treated dogs (5th stage suceptible instars) USED

fit data with logistic curve -using % dead 5th stage suceptible bugs against days post fluralaner tx -fit2 stores fitted values -plot to check fit -extract fit values using equation y = Asym / (1 + exp((xmid - input) / scal))

```
#Initial vectors for days post treatment, % killed
x <- c(4, 30, 60, 90, 120, 210, 360) #days post treatment
y <- c(1.0, 1.0, 0.99, 0.49, 0.47, 0, 0)  #% killed
z <- c(1.0, 0.99, 0.99, 0.79, 0.7, 0.02, 0) #% killed

fit2 <- nls(z ~ SSlogis(x, Asym, xmid, scal), data = data.frame(x, z))
Asym<-summary(fit2)$parameters[1,1]
xmid<-summary(fit2)$parameters[2,1]
scal<-summary(fit2)$parameters[3,1]
```

**Parameters** Y - the proportion of triatomines infected  
X - the proportion of dogs infected  
a is expected number of bites on dogs per triatomine (1/14)  
m is equilibrium triatomine density per dog  
n is length of incubation period in insects (45)  
g is the daily force of triatomine mortality, mzl at K, the carrying capacity  
b is the transmission efficiency from infectious triatomines to susceptible dog, through biting (0.00068)  
c is the probability of infection of an uninfected triatomine by biting an infectious dog  
k is the probabilty of tranmission through oral ingestion of vectors (0.1)  
r is daily force of infected dog mortality: 1.38/1460= 0.0009  
p is probability dog eats the bug (to be varied)  
R is maximum birthrate (estimate from T. brasiliensis)  
K is carrying capacity of vectors per dog

force the time dependent covariate  
z is the probability of death from Fluralaner per bite  
jr added code to apply more than one treatment  
jr added function for implementing treatments at different times

**Treatment model**

```
trt2.function <- function(fit2, trt.days, signal) {
  Asym<-summary(fit2)$parameters[1,1]
  xmid<-summary(fit2)$parameters[2,1]
  scal<-summary(fit2)$parameters[3,1]
  trt.start <- trt.days
  trt.end <- c(trt.start + 400)
  trt.segments <- unlist(Map(':', trt.start, trt.end))
  time.segments <- rep(0:401, times=length(trt.start)) 
  signal$import[trt.segments] = (Asym / (1 + exp((xmid - times[time.segments]) / scal)))  
  return(signal)
}

RMTx2 <- function(times, stateTx2, parametersTx2)   
{
  with(
    as.list(c(stateTx2, parametersTx2)), 
    {
      z <- input(times)
      dX <- (((m*a*b*Y)+MM)+(p*k*(a*m*z*Y)))*(1-X)-r*X 
      dY <- a*c*X*(exp(-g*n)-Y)-((g*Y)+(m*a*z*Y))
      dm <- (R*(1-m/K)*m )+(-m*a*z)
      return(list(c(dX, dY, dm)))
    }
  )
}
```

#### Paper figures

S1

```
df1 <- data.frame(X=x, Z=z)
df2 <- data.frame(time = c(0:365))%>%
  mutate(prop = Asym/(1+exp((xmid-time)/scal)))

ggplot()+
  theme_classic()+
  geom_line(data=df2, aes(x=time, y=prop), size=2, color= "gray")+
  geom_point(data=df1, aes(x=X, y=Z), size=5, shape=8, color="black")+
  theme(text = element_text(size = 20))+    
  xlab("Days Post-Treatment") + ylab("Proportion killed")
```

##### High prevalence, domestic vectors

S2A

```
#set parameters and run model
init <- c(X = 0.01, Y = 0, m= 40)
parameters <- c(a=1/14, b=0.00068, m=10, n=45, g=0.005, c=0.28, r=1/(3*365), K = 40, R= 0.09)
times <- seq(0, 40000, by = 1)
out <- as.data.frame(ode(y = init, times = times, func = RM, parms = parameters))%>%
  mutate(year = times/365.25)

pal1 <- c('Infected dogs' = "red3",
          'Infected bugs' = "dodgerblue3")

#Extract equilibrium values
tail(out)
```

```
##        time         X         Y  m     year
## 39996 39995 0.5368288 0.5448028 40 109.5003
## 39997 39996 0.5368288 0.5448028 40 109.5031
## 39998 39997 0.5368288 0.5448028 40 109.5058
## 39999 39998 0.5368288 0.5448028 40 109.5086
## 40000 39999 0.5368288 0.5448028 40 109.5113
## 40001 40000 0.5368288 0.5448028 40 109.5140
```

```
#plot
ggplot()+
  theme_classic()+
  geom_line(data=out, aes(x=year, y=X, color="Infected dogs"), size=2)+
  geom_line(data=out, aes(x=year, y=Y, color="Infected bugs"), size=2)+
  scale_color_manual(values = pal1, name= "Disease state")+ 
  scale_x_continuous(limits = c(0, 40))+
  theme(text = element_text(size = 20))+                    
  xlab("Time (years)") + ylab("Proportion of population")
```

Figure 2

```
#set up time scale
times <- seq(0, 35000, by = 1)

#Extract equilibrium
RMequil <- getEq(a=1/14, b=.00068, m=40, n=45, g=.005, c=.28, r=1/(3*365))
Xeq <- RMequil[2]
Yeq <- RMequil[3]

signal <- data.frame(times = times, import = rep(0, length(times)))
trt.days <- c(10000) 
out <-trt2.function(fit2, trt.days, signal)
input <- approxfun(out, rule = 2)

#Set params/ init conditions
initTx2 <- c(X = Xeq, Y= Yeq, m=40) 
parametersTx2 <- c(a=1/14, b=0.00068, m=40, n=45, g=0.005, c=0.28, k=.10, r= 1/(3*365), p=0.8, K=40, R= 0.09, MM= 0)

#Run model
outTx2 <- as.data.frame(ode(y = initTx2, times = times, func = RMTx2, parms = parametersTx2))%>%
  mutate(year=times/365.25)

#plot
ggplot()+
  theme_classic()+
  geom_line(data=outTx2, aes(x=year, y=X, color="Infected dogs"), size=2)+
  geom_line(data=outTx2, aes(x=year, y=Y, color="Infected bugs"), size=2)+
  geom_vline(data = as.data.frame(trt.days) %>% mutate(trt.days = trt.days/365.25),  
             aes(xintercept = trt.days),   
             linetype =  "twodash", color = "black", size = 1)+
  scale_color_manual(values = pal1, name= "Disease state")+ 
  scale_x_continuous(limits = c(25, 40))+
  theme(text = element_text(size = 20))+                    
  xlab("Time (years)") + ylab("Proportion of population")
```

Figure 3A: One treatment/yr x 4yrs

```
#Extract equilibrium
trt.days <- seq(10000, 10000+365.25*3, 365.25) 
out <-trt2.function(fit2, trt.days, signal)
input <- approxfun(out, rule = 2)

#Run model
outTx2 <- as.data.frame(ode(y = initTx2, times = times, func = RMTx2, parms = parametersTx2))%>%
  mutate(year=times/365.25)

#plot
ggplot()+
  theme_classic()+
  geom_line(data=outTx2, aes(x=year, y=X, color="Infected dogs"), size=2)+
  geom_line(data=outTx2, aes(x=year, y=Y, color="Infected bugs"), size=2)+
  geom_vline(data = as.data.frame(trt.days) %>% mutate(trt.days = trt.days/365.25),  
             aes(xintercept = trt.days),   
             linetype =  "twodash", color = "black", size = 1)+
  scale_color_manual(values = pal1, name= "Disease state")+ 
  scale_x_continuous(limits = c(25, 40))+
  theme(text = element_text(size = 20))+                    
  xlab("Time (years)") + ylab("Proportion of population")
```

Figure 3B: One treatment/yr x 6yrs

```
#Extract equilibrium
trt.days <- seq(10000, 10000+365.25*5, 365.25) 
out <-trt2.function(fit2, trt.days, signal)
input <- approxfun(out, rule = 2)

#Run model
outTx2 <- as.data.frame(ode(y = initTx2, times = times, func = RMTx2, parms = parametersTx2))%>%
  mutate(year=times/365.25)

#plot
fig <- ggplot()+
  theme_classic()+
  geom_line(data=outTx2, aes(x=year, y=X, color="Infected dogs"), size=2)+
  geom_line(data=outTx2, aes(x=year, y=Y, color="Infected bugs"), size=2)+
  geom_vline(data = as.data.frame(trt.days) %>% mutate(trt.days = trt.days/365.25),  
             aes(xintercept = trt.days),   
             linetype =  "twodash", color = "black", size = 1)+
  scale_color_manual(values = pal1, name= "Disease state")+ 
  scale_x_continuous(limits = c(25, 40))+
  theme(text = element_text(size = 20))+                    
  xlab("Time (years)") + ylab("Proportion of population")
fig
```

Figure 3C: Treatment every 90 days for a year

```
#Extract equilibrium
trt.days <- seq(10000, 10359, 90) 
out <-trt2.function(fit2, trt.days, signal)
input <- approxfun(out, rule = 2)

#Run model
outTx2 <- as.data.frame(ode(y = initTx2, times = times, func = RMTx2, parms = parametersTx2))%>%
  mutate(year=times/365.25)

#plot
ggplot()+
  theme_classic()+
  geom_line(data=outTx2, aes(x=year, y=X, color="Infected dogs"), size=2)+
  geom_line(data=outTx2, aes(x=year, y=Y, color="Infected bugs"), size=2)+
  geom_vline(data = as.data.frame(trt.days) %>% mutate(trt.days = trt.days/365.25),  
             aes(xintercept = trt.days),   
             linetype =  "twodash", color = "black", size = 1)+
  scale_color_manual(values = pal1, name= "Disease state")+ 
  scale_x_continuous(limits = c(25, 40))+
  theme(text = element_text(size = 20))+                    
  xlab("Time (years)") + ylab("Proportion of population")
```

Figure 3D: Treatment every 90 days for 2 years

```
#Extract equilibrium
trt.days <- seq(10000, 10000+360*2-1, 90) 
out <-trt2.function(fit2, trt.days, signal)
input <- approxfun(out, rule = 2)

#Run model
outTx2 <- as.data.frame(ode(y = initTx2, times = times, func = RMTx2, parms = parametersTx2))%>%
  mutate(year=times/365.25)

#plot
ggplot()+
  theme_classic()+
  geom_line(data=outTx2, aes(x=year, y=X, color="Infected dogs"), size=2)+
  geom_line(data=outTx2, aes(x=year, y=Y, color="Infected bugs"), size=2)+
  geom_vline(data = as.data.frame(trt.days) %>% mutate(trt.days = trt.days/365.25),  
             aes(xintercept = trt.days),   
             linetype =  "twodash", color = "black", size = 1)+
  scale_color_manual(values = pal1, name= "Disease state")+ 
  scale_x_continuous(limits = c(25, 40))+
  theme(text = element_text(size = 20))+                    
  xlab("Time (years)") + ylab("Proportion of population")
```

##### Low prevalence, sylvatic vectors

**S2B: Low Disease prevalence, sylvatic vectors**

```
#set parameters and run model
init <- c(X = 0.01, Y = 0, m= 15)
parameters <- c(a=1/14, b=0.00068, n=45, g=0.005, c=0.28, r=1/(3*365), K = 15, R= 0.09)
times <- seq(0, 20000, by = 1)
out <- as.data.frame(ode(y = init, times = times, func = RM, parms = parameters))%>%
  mutate(year = times/365.25)

#Extract equilibrium values
tail(out)
```

```
##        time         X         Y  m     year
## 19996 19995 0.2364268 0.3881171 15 54.74333
## 19997 19996 0.2364268 0.3881171 15 54.74606
## 19998 19997 0.2364268 0.3881171 15 54.74880
## 19999 19998 0.2364268 0.3881171 15 54.75154
## 20000 19999 0.2364268 0.3881171 15 54.75428
## 20001 20000 0.2364268 0.3881171 15 54.75702
```

```
#plot results
ggplot()+
  theme_classic()+
  geom_line(data=out, aes(x=year, y=X, color="Infected dogs"), size=2)+
  geom_line(data=out, aes(x=year, y=Y, color="Infected bugs"), size=2)+
  scale_color_manual(values = pal1, name= "Disease state")+ 
  scale_x_continuous(limits = c(0, 40))+
  theme(text = element_text(size = 20))+                    
  xlab("Time (years)") + ylab("Proportion of population")
```

Figure 4A: One treatment, life span = 3yr

```
#set up time scale
times <- seq(0, 35000, by = 1)

#Extract equilibrium
RMequil <- getEq(a=1/14, b=.00068, m=40, n=45, g=.005, c=.28, r=1/(3*365))
Xeq <- RMequil[2]
Yeq <- RMequil[3]

signal <- data.frame(times = times, import = rep(0, length(times)))
trt.days <- c(10000) 
out <-trt2.function(fit2, trt.days, signal)
input <- approxfun(out, rule = 2)

#Set params/ init conditions
initTx2 <- c(X = Xeq, Y= Yeq, m=40)
parametersTx2 <- c(a=1/14, b=0.00068, m=40, n=45, g=0.005, c=0.28, k=.10, r= 1/(3*365), p=0.8, K=15, R= 0.09, MM= 0)


parameters <- c(a=1/14, b=0.00068, n=45, g=0.005, c=0.28, r=1/(3*365), K = 15, R= 0.09)

#Run model
outTx2 <- as.data.frame(ode(y = initTx2, times = times, func = RMTx2, parms = parametersTx2))%>%
  mutate(year=times/365.25)

#plot
ggplot()+
  theme_classic()+
  geom_line(data=outTx2, aes(x=year, y=X, color="Infected dogs"), size=2)+
  geom_line(data=outTx2, aes(x=year, y=Y, color="Infected bugs"), size=2)+
  geom_vline(data = as.data.frame(trt.days) %>% mutate(trt.days = trt.days/365.25),  
             aes(xintercept = trt.days),   
             linetype =  "twodash", color = "black", size = 1)+
  scale_color_manual(values = pal1, name= "Disease state")+ 
  scale_x_continuous(limits = c(25, 40))+
  theme(text = element_text(size = 20))+                    
  xlab("Time (years)") + ylab("Proportion of population")
```

Figure 4B: One treatment/yr x 4yrs, life span = 3yr

```
#Extract equilibrium
trt.days <- seq(10000, 10000+365.25*3, 365.25) 
out <-trt2.function(fit2, trt.days, signal)
input <- approxfun(out, rule = 2)

#Run model
outTx2 <- as.data.frame(ode(y = initTx2, times = times, func = RMTx2, parms = parametersTx2))%>%
  mutate(year=times/365.25)

#plot
ggplot()+
  theme_classic()+
  geom_line(data=outTx2, aes(x=year, y=X, color="Infected dogs"), size=2)+
  geom_line(data=outTx2, aes(x=year, y=Y, color="Infected bugs"), size=2)+
  geom_vline(data = as.data.frame(trt.days) %>% mutate(trt.days = trt.days/365.25),  
             aes(xintercept = trt.days),   
             linetype =  "twodash", color = "black", size = 1)+
  scale_color_manual(values = pal1, name= "Disease state")+ 
  scale_x_continuous(limits = c(25, 40))+
  theme(text = element_text(size = 20))+                    
  xlab("Time (years)") + ylab("Proportion of population")
```

Figure 4C: Treatment every 90 days for a year, life span = 3yr

```
#Extract equilibrium
trt.days <- seq(10000, 10359, 90) 
out <-trt2.function(fit2, trt.days, signal)
input <- approxfun(out, rule = 2)

#Run model
outTx2 <- as.data.frame(ode(y = initTx2, times = times, func = RMTx2, parms = parametersTx2))%>%
  mutate(year=times/365.25)

#plot
ggplot()+
  theme_classic()+
  geom_line(data=outTx2, aes(x=year, y=X, color="Infected dogs"), size=2)+
  geom_line(data=outTx2, aes(x=year, y=Y, color="Infected bugs"), size=2)+
  geom_vline(data = as.data.frame(trt.days) %>% mutate(trt.days = trt.days/365.25),  
             aes(xintercept = trt.days),   
             linetype =  "twodash", color = "black", size = 1)+
  scale_color_manual(values = pal1, name= "Disease state")+ 
  scale_x_continuous(limits = c(25, 40))+
  theme(text = element_text(size = 20))+                    
  xlab("Time (years)") + ylab("Proportion of population")
```

Figure 4D: One treatment, life span = 6yr

```
#set up time scale
times <- seq(0, 35000, by = 1)

#Extract equilibrium
RMequil <- getEq(a=1/14, b=.00068, m=40, n=45, g=.005, c=.28, r=1/(6*365))
Xeq <- RMequil[2]
Yeq <- RMequil[3]

signal <- data.frame(times = times, import = rep(0, length(times)))
trt.days <- c(10000) 
out <-trt2.function(fit2, trt.days, signal)
input <- approxfun(out, rule = 2)

#Set params/ init conditions
initTx2 <- c(X = Xeq, Y= Yeq, m=40)

# parametersTx2 <- c(a=1/14, b=0.00068, m=40, n=45, g=0.005, c=0.28, k=.10, r= 1/(6*365), p=0.8, K=15, R= 0.09, MM= 0)

parametersTx2 <- c(a=1/14, b=0.00068, m=40, n=45, g=0.005, c=0.28, k=.10, r= 1/(6*365), p=0.8, K=7, R= 0.09, MM= 0)

#Run model
outTx2 <- as.data.frame(ode(y = initTx2, times = times, func = RMTx2, parms = parametersTx2))%>%
  mutate(year=times/365.25)

#plot
ggplot()+
  theme_classic()+
  geom_line(data=outTx2, aes(x=year, y=X, color="Infected dogs"), size=2)+
  geom_line(data=outTx2, aes(x=year, y=Y, color="Infected bugs"), size=2)+
  geom_vline(data = as.data.frame(trt.days) %>% mutate(trt.days = trt.days/365.25),  
             aes(xintercept = trt.days),   
             linetype =  "twodash", color = "black", size = 1)+
  scale_color_manual(values = pal1, name= "Disease state")+ 
  scale_x_continuous(limits = c(25, 40))+
  theme(text = element_text(size = 20))+                    
  xlab("Time (years)") + ylab("Proportion of population")
```

Figure 4E: One treatment/yr x 4yrs, life span = 6yr

```
#Extract equilibrium
trt.days <- seq(10000, 10000+365.25*3, 365.25) 
out <-trt2.function(fit2, trt.days, signal)
input <- approxfun(out, rule = 2)

#Run model
outTx2 <- as.data.frame(ode(y = initTx2, times = times, func = RMTx2, parms = parametersTx2))%>%
  mutate(year=times/365.25)

#plot
ggplot()+
  theme_classic()+
  geom_line(data=outTx2, aes(x=year, y=X, color="Infected dogs"), size=2)+
  geom_line(data=outTx2, aes(x=year, y=Y, color="Infected bugs"), size=2)+
  geom_vline(data = as.data.frame(trt.days) %>% mutate(trt.days = trt.days/365.25),  
             aes(xintercept = trt.days),   
             linetype =  "twodash", color = "black", size = 1)+
  scale_color_manual(values = pal1, name= "Disease state")+ 
  scale_x_continuous(limits = c(25, 40))+
  theme(text = element_text(size = 20))+                    
  xlab("Time (years)") + ylab("Proportion of population")
```

Figure 4F: Treatment every 90 days for a year, life span = 6yr

```
#Extract equilibrium
trt.days <- seq(10000, 10359, 90) 
out <-trt2.function(fit2, trt.days, signal)
input <- approxfun(out, rule = 2)

#Run model
outTx2 <- as.data.frame(ode(y = initTx2, times = times, func = RMTx2, parms = parametersTx2))%>%
  mutate(year=times/365.25)

#plot
ggplot()+
  theme_classic()+
  geom_line(data=outTx2, aes(x=year, y=X, color="Infected dogs"), size=2)+
  geom_line(data=outTx2, aes(x=year, y=Y, color="Infected bugs"), size=2)+
  geom_vline(data = as.data.frame(trt.days) %>% mutate(trt.days = trt.days/365.25),  
             aes(xintercept = trt.days),   
             linetype =  "twodash", color = "black", size = 1)+
  scale_color_manual(values = pal1, name= "Disease state")+ 
  scale_x_continuous(limits = c(25, 40))+
  theme(text = element_text(size = 20))+                    
  xlab("Time (years)") + ylab("Proportion of population")
```

##### Low prevalence, sylvatic vectors + semi-sylvatic vactors

Figure 5A: One treatment, life span = 3yr

```
#set up time scale
times <- seq(0, 35000, by = 1)

#Extract equilibrium
RMequil <- getEq(a=1/14, b=.00068, m=40, n=45, g=.005, c=.28, r=1/(3*365))
Xeq <- RMequil[2]
Yeq <- RMequil[3]

signal <- data.frame(times = times, import = rep(0, length(times)))
trt.days <- c(10000) 
out <-trt2.function(fit2, trt.days, signal)
input <- approxfun(out, rule = 2)

#Set params/ init conditions
initTx2 <- c(X = Xeq, Y= Yeq, m=40)
parametersTx2 <- c(a=1/14, b=0.00068, m=40, n=45, g=0.005, c=0.28, k=.10, r= 1/(3*365), p=0.8, K=15, R= 0.09, MM= 0.0001)


parameters <- c(a=1/14, b=0.00068, n=45, g=0.005, c=0.28, r=1/(3*365), K = 15, R= 0.09)

#Run model
outTx2 <- as.data.frame(ode(y = initTx2, times = times, func = RMTx2, parms = parametersTx2))%>%
  mutate(year=times/365.25)

#plot
ggplot()+
  theme_classic()+
  geom_line(data=outTx2, aes(x=year, y=X, color="Infected dogs"), size=2)+
  geom_line(data=outTx2, aes(x=year, y=Y, color="Infected bugs"), size=2)+
  geom_vline(data = as.data.frame(trt.days) %>% mutate(trt.days = trt.days/365.25),  
             aes(xintercept = trt.days),   
             linetype =  "twodash", color = "black", size = 1)+
  scale_color_manual(values = pal1, name= "Disease state")+ 
  scale_x_continuous(limits = c(25, 40))+
  theme(text = element_text(size = 20))+                    
  xlab("Time (years)") + ylab("Proportion of population")
```

Figure 5B: One treatment/yr x 4yrs, life span = 3yr

```
#Extract equilibrium
trt.days <- seq(10000, 10000+365.25*3, 365.25) 
out <-trt2.function(fit2, trt.days, signal)
input <- approxfun(out, rule = 2)

#Run model
outTx2 <- as.data.frame(ode(y = initTx2, times = times, func = RMTx2, parms = parametersTx2))%>%
  mutate(year=times/365.25)

#plot
ggplot()+
  theme_classic()+
  geom_line(data=outTx2, aes(x=year, y=X, color="Infected dogs"), size=2)+
  geom_line(data=outTx2, aes(x=year, y=Y, color="Infected bugs"), size=2)+
  geom_vline(data = as.data.frame(trt.days) %>% mutate(trt.days = trt.days/365.25),  
             aes(xintercept = trt.days),   
             linetype =  "twodash", color = "black", size = 1)+
  scale_color_manual(values = pal1, name= "Disease state")+ 
  scale_x_continuous(limits = c(25, 40))+
  theme(text = element_text(size = 20))+                    
  xlab("Time (years)") + ylab("Proportion of population")
```

Figure 4C: Treatment every 90 days for a year, life span = 3yr

```
#Extract equilibrium
trt.days <- seq(10000, 10359, 90) 
out <-trt2.function(fit2, trt.days, signal)
input <- approxfun(out, rule = 2)

#Run model
outTx2 <- as.data.frame(ode(y = initTx2, times = times, func = RMTx2, parms = parametersTx2))%>%
  mutate(year=times/365.25)

#plot
ggplot()+
  theme_classic()+
  geom_line(data=outTx2, aes(x=year, y=X, color="Infected dogs"), size=2)+
  geom_line(data=outTx2, aes(x=year, y=Y, color="Infected bugs"), size=2)+
  geom_vline(data = as.data.frame(trt.days) %>% mutate(trt.days = trt.days/365.25),  
             aes(xintercept = trt.days),   
             linetype =  "twodash", color = "black", size = 1)+
  scale_color_manual(values = pal1, name= "Disease state")+ 
  scale_x_continuous(limits = c(25, 40))+
  theme(text = element_text(size = 20))+                    
  xlab("Time (years)") + ylab("Proportion of population")
```

Figure 5D: One treatment, life span = 6yr

```
#set up time scale
times <- seq(0, 35000, by = 1)

#Extract equilibrium
RMequil <- getEq(a=1/14, b=.00068, m=40, n=45, g=.005, c=.28, r=1/(6*365))
Xeq <- RMequil[2]
Yeq <- RMequil[3]

signal <- data.frame(times = times, import = rep(0, length(times)))
trt.days <- c(10000) 
out <-trt2.function(fit2, trt.days, signal)
input <- approxfun(out, rule = 2)

#Set params/ init conditions
initTx2 <- c(X = Xeq, Y= Yeq, m=40)

parametersTx2 <- c(a=1/14, b=0.00068, m=40, n=45, g=0.005, c=0.28, k=.10, r= 1/(6*365), p=0.8, K=7, R= 0.09, MM= 0.0001)

#Run model
outTx2 <- as.data.frame(ode(y = initTx2, times = times, func = RMTx2, parms = parametersTx2))%>%
  mutate(year=times/365.25)

#plot
ggplot()+
  theme_classic()+
  geom_line(data=outTx2, aes(x=year, y=X, color="Infected dogs"), size=2)+
  geom_line(data=outTx2, aes(x=year, y=Y, color="Infected bugs"), size=2)+
  geom_vline(data = as.data.frame(trt.days) %>% mutate(trt.days = trt.days/365.25),  
             aes(xintercept = trt.days),   
             linetype =  "twodash", color = "black", size = 1)+
  scale_color_manual(values = pal1, name= "Disease state")+ 
  scale_x_continuous(limits = c(25, 40))+
  theme(text = element_text(size = 20))+                    
  xlab("Time (years)") + ylab("Proportion of population")
```

Figure 5E: One treatment/yr x 4yrs, life span = 3yr

```
#Extract equilibrium
trt.days <- seq(10000, 10000+365.25*3, 365.25) 
out <-trt2.function(fit2, trt.days, signal)
input <- approxfun(out, rule = 2)

#Run model
outTx2 <- as.data.frame(ode(y = initTx2, times = times, func = RMTx2, parms = parametersTx2))%>%
  mutate(year=times/365.25)

#plot
ggplot()+
  theme_classic()+
  geom_line(data=outTx2, aes(x=year, y=X, color="Infected dogs"), size=2)+
  geom_line(data=outTx2, aes(x=year, y=Y, color="Infected bugs"), size=2)+
  geom_vline(data = as.data.frame(trt.days) %>% mutate(trt.days = trt.days/365.25),  
             aes(xintercept = trt.days),   
             linetype =  "twodash", color = "black", size = 1)+
  scale_color_manual(values = pal1, name= "Disease state")+ 
  scale_x_continuous(limits = c(25, 40))+
  theme(text = element_text(size = 20))+                    
  xlab("Time (years)") + ylab("Proportion of population")
```

Figure 5F: Treatment every 90 days for a year, life span = 3yr

```
#Extract equilibrium
trt.days <- seq(10000, 10359, 90) 
out <-trt2.function(fit2, trt.days, signal)
input <- approxfun(out, rule = 2)

#Run model
outTx2 <- as.data.frame(ode(y = initTx2, times = times, func = RMTx2, parms = parametersTx2))%>%
  mutate(year=times/365.25)

#plot
ggplot()+
  theme_classic()+
  geom_line(data=outTx2, aes(x=year, y=X, color="Infected dogs"), size=2)+
  geom_line(data=outTx2, aes(x=year, y=Y, color="Infected bugs"), size=2)+
  geom_vline(data = as.data.frame(trt.days) %>% mutate(trt.days = trt.days/365.25),  
             aes(xintercept = trt.days),   
             linetype =  "twodash", color = "black", size = 1)+
  scale_color_manual(values = pal1, name= "Disease state")+ 
  scale_x_continuous(limits = c(25, 40))+
  theme(text = element_text(size = 20))+                    
  xlab("Time (years)") + ylab("Proportion of population")
```
